# Seroprevalence and associated risk factors of *Toxoplasma gondii* in sheep and goats of mid-western Nepal with potential risk of human infections

**DOI:** 10.1101/2025.06.07.25328778

**Authors:** Bhrikuti Bhattarai Sharma, Ananta Dahal, Rebanta Kumar Bhattarai, Jaya Prasad Singh, Prativa Shrestha, Anup Adhikari, Deb Prasad Pandey

## Abstract

**Introduction:** *Toxoplasma gondii* is a protozoan that causes zoonotic disease in warm-blooded animals including humans worldwide. We aimed to evaluate the seroprevalence of *T. gondii* infection and associated risk factors in sheep and goats and analyze risk to farm attendants across three different eco-zones in mid-western Nepal.

**Methods:** We conducted a cross-sectional study of 368 sheep and goats in three districts representing distinct eco-zones and evaluated the seroprevalence by detecting *T. gondii* antibodies withan indirect enzyme-linked immunosorbent assay. Chi-square test was used to assess potential risk factors while spearman’s rank correlation was used for risk assessment in farm attandants.

**Results:** The herd-level and individual animal level seroprevalence of *T. gondii* infection was 92.8% (n = 64/69) and 61.7% (n = 227/368), respectively. Sheep had higher seroprevalence (81.4%, n = 136/167) than goats (45.3%, n = 91/201). Altitudinal gradient had a weak but positive correlation to the herd seroprevalence (r = 0.38). The high seroprevalence suggests that *T. gondii* is widespread in the study area and sheep and goats could serve as significant sources of transmission to humans.

**Conclusion:** Altitude, age of animal, type of host animal, herd size, rearing system, types of animals reared in a herd, presence of domestic cats and access of cats to water sources were detected as potential risk factors for *T. gondii* infection in sheep and goats. There is a need for additional epidemiological studies and interventions for appropriate prevention and control strategies for *T. gondii* infections in sheep, goats, and humans.

**Author summary:** Toxoplasmosis, caused by *Toxoplasma gondii*, is a widespread zoonotic disease that affects a wide range of warm-blooded animals, including humans. We evaluated the seroprevalence of *T. gondii* infection and its associated risk factors in sheep and goats across three eco-zones in mid-western Nepal. A total of 368 animals from three districts were included in this cross-sectional study. We found a high herd-level seroprevalence of 92.8%, with an overall individual animal seroprevalence of 61.7%. Sheep had a significantly higher seroprevalence (81.4%) compared to goats (45.3%). We also identified several risk factors associated with *T. gondii* infection, including altitude, animal age, host type, herd size, rearing system, and the presence of domestic cats. In addition, we assessed the behaviors of farm attendants, revealing a moderate correlation with the seroprevalence of *T. gondii* at the herd level. The high seroprevalence in sheep and goats suggests that these animals may play a key role in the transmission of *T. gondii* to humans, particularly in rural areas with close livestock-human interactions. The study highlights the need for additional epidemiological research and the development of targeted prevention and control strategies for *T. gondii* in livestock populations to reduce the risk of zoonotic transmission.

## Introduction

*Toxoplasma gondii* (Family: Sarcocystidae, Sub-family: Toxoplasmatinae) is a cyst-forming apicomplexan, obligate intracellular protozoan parasite that causes toxoplasmosis in warm-blooded animals worldwide [1]. This coccidian, zoonotic parasite opportunistically infects these hosts in which it can persist for long period. Cats (domesticated, feral and wild) and other felids (e.g., bobcat, tiger, leopard) are the definitive hosts in which oocysts are developed in their small intestine. In optimum environmental conditions (temperature and oxygen), the released oocysts develop sporozoites within one to five days [2]. Other warm-blooded animals (e.g., sheep, goats, rodents, birds, etc.) including humans are the intermediate hosts which are infected with either sporulated oocysts that release sporozoites or with tissue cysts that release bradyzoites into stomach. These sporozoites and bradyzoites cross intestinal lining and develop into tachyzoites that travel throughout the body. These three stages (sporozoites, tachyzoites and bradyzoites) are infective to definitive and intermediate hosts [2].

The intermediate hosts can get infection through the consumption of unpasteurized milk, contaminated foods with sporulated oocysts, and raw or undercooked meat containing bradyzoites or tachyzoites and through the placenta [3–5]. Cats typically get infected with *T. gondii* by hunting and consuming meat of infected intermediate hosts. Cats barely show the clinical signs even during the shedding of oocysts i.e., sub-clinical form occurs in cats. However, cats play an important role in the transmission of toxoplasmosis [6]. The consumption of sheep and goat meat is considered one of the major transmission routes for *T. gondii* infection in humans [7]. Most of the humans infected with Toxoplasma, have no signs and symptoms. However, some people show signs and symptoms similar to those of the flu, inclusive of body aches, swollen lymph nodes, headache, fever, fatigue, confusion, poor coordination, and seizures. Toxoplasma infection in humans can cause major harm, particularly to those with weakened immune systems, HIV/AIDS patients, pregnant mothers, young children, and those having other medical conditions [8]. Both congenital infection of the fetus and abortion are possible outcomes of infection in pregnant women [9,10]. Retinochoroiditis, hydrocephalus, and intracranial calcification are the outcomes of fetal infections occurring at birth in humans [11]. Janku, a Prague-based ophthalmologist, reported the first case of toxoplasmosis cyst in the retina of a child [11]. Nearly one third of the world human population are infected by *T. gondii* [12]. In Nepal, the first case of congenital toxoplasmosis was detected in a 53 days baby with hepatosplenomegaly and optic nerve colobema with a large scar in the right eye [13]. Nepal has listed toxoplasmosis as one of the top ten zoonotic diseases [14].

Globally, sheep, goats, pigs, cattle, poultry, and cats are the most seriously affected domestic animals by *T. gondii* [15]. Grazing sheep and goats can get infected while grazing through the contaminated pasture. Evidence of the presence of *Toxoplasma* in the milk of sheep and goat, can also be a route for transmission of this parasite [16]. Its infection causes low productivity leading to unexpected loss of milk and meat and threatening food safety widely. *T. gondii* was a significant contributor to sheep abortions in New Zealand [17]. Toxoplasmosis was diagnosed in aborted sheep and goats in Egypt [18], Jordan [19] and Ethiopia [20]. Overall, significant illnesses have been reported worldwide with varying degrees of illness in humans and animals [21]. Therefore, this parasite has significant importance in the medical and veterinary sectors. However, there is a scarcity of information about the status of toxoplasmosis in sheep and goats and possible infections to associated farmers in Nepal.

Few studies on toxoplasmosis in sheep and goats of Nepal had been carried out [22,23–25]. Although these investigations showed sheep and goats toxoplasmosis, they did not concentrate on the prevalence and risk factors of *T. gondii* in sheep and goats across the altitudinal gradient in Nepal. Given the lack of research on toxoplasmosis throughout the nation as well as across its diverse altitudes and climatic zones, additional studies should be conducted across altitudinal gradients in this country to understand a wide range of risk factors. Due to their higher risk of disease and substantial contribution to meat consumption in different regions of Nepal, small ruminants are given priority. Because raw or undercooked mutton and chevon may carry toxoplasmosis to humans and other animals, we aimed to determine the seroprevalence and associated risk factors of *T. gondii* infection in sheep and goats and identify risks of *T. gondii* infection among vulnerable farm attendants across altitudinal gradients of different ecozones in mid-western Nepal. Our findings provide information required to formulate measures for the prevention and control of toxoplasmosis in Nepalese communities and elsewhere having similar socio-economic and geo-climatic conditions.

## Methods

### Ethical Statement

We interviewed farm attendants face-to-face after the farm owners gave their verbal permission and then we sampled sheep and goats after taking permission from farm owners We have not presented any identifiable image of farm attendants in our study. This study was approved by the Institutional Review Board of AFU, Chitwan, Nepal (Ref. no. AFU-FW A00031653).

### Study area and populations

#### Study area

Mid-western part of Nepal consists of Karnali Province and some parts of Lumbini Province. This region has three topographically divided ecological regions: Mountains, Hills and Terai. The climate varies from alpine in the mountains, temperate in the hills and tropical in the terai areas of the mid-western region of Nepal. Based on the availability of sheep and goat farms, we conducted the study in three purposively selected Districts (Jumla, Surkhet and Banke) representing those ecological regions, respectively (Figure 1).

**Figure 1.**
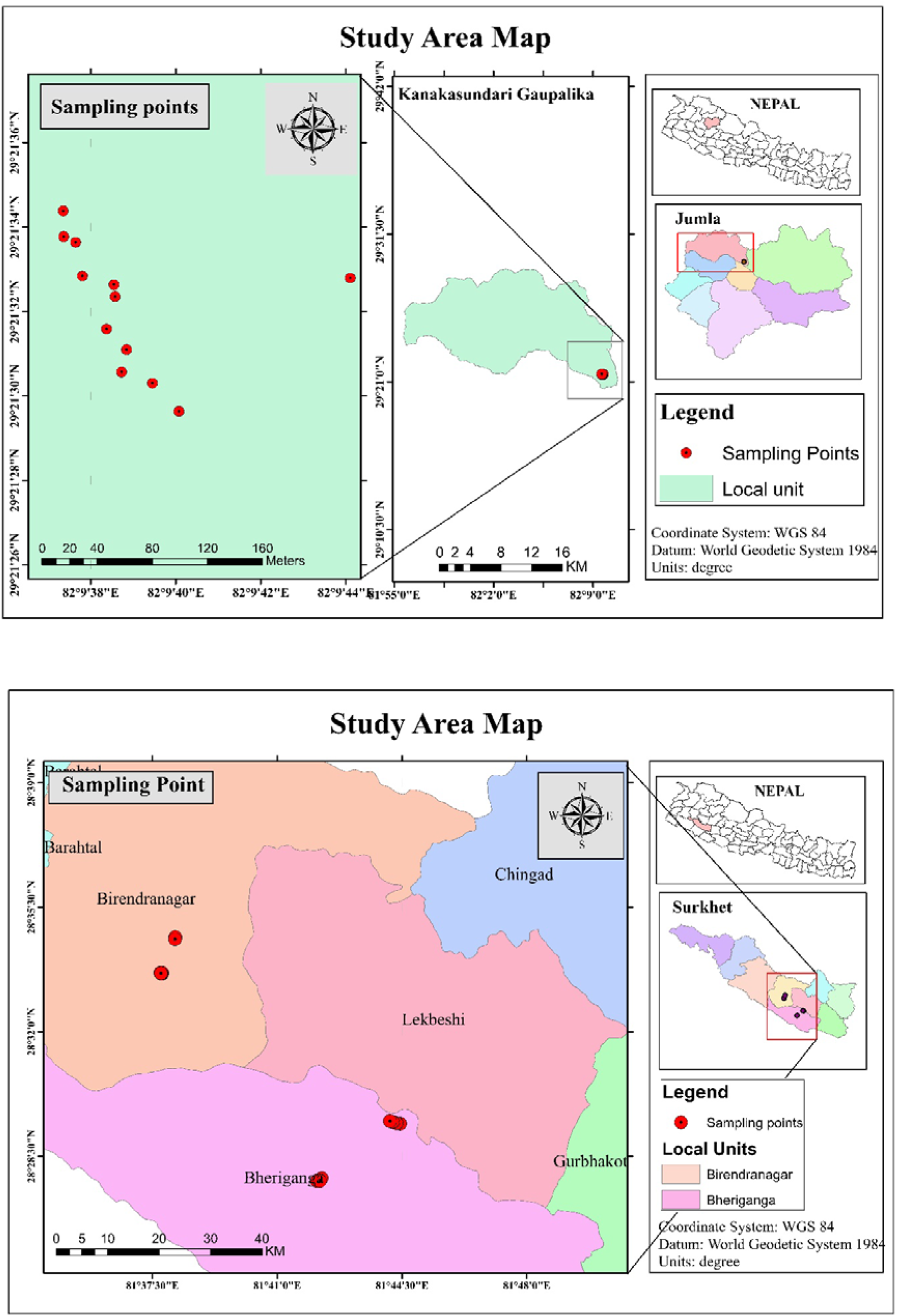
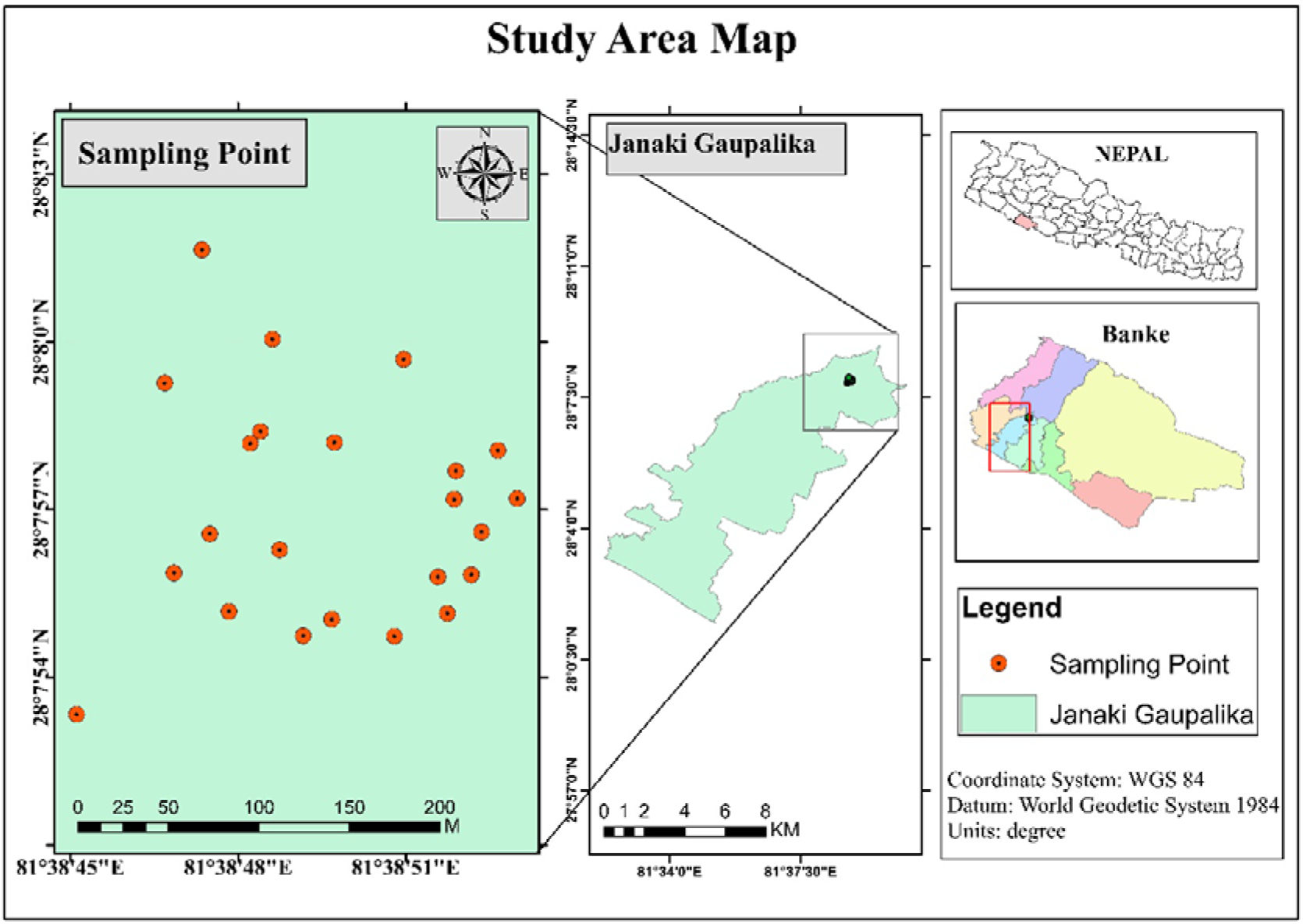
Map of study districts: Jumla, Surkhet and Banke from top to bottom respectively.

Jumla District is a mountainous District elevated from 915 meters to 4,679 meters above sea level (masl). It is extended in 2,531 square kilometers (sq. km.) across a longitude of 81° 28’ to 82° 18’ East and latitude of 28° 58’ to 29° 30’ North and has temperature ranging from 18–30 °C in summer and minus 14–8 °C in winter with average rainfall of 1,343 mm [26]. It is one of the remote and backward districts of Karnali Province [27] and has 24,438 households representing 118,349 human populations (S1 Table).

Surkhet, the capital of Karnali Province is a hilly District elevated from 198 to 2,367 masl and extended in 2,451 sq. km. across a longitude of 80° 59’ to 82° 02’ E and latitude of 28° 20’ to 28° 58’ N. The temperature of this District ranges from 5–38 °C in winter and summer, respectively [28]. This District has 97,893 households representing 415,126 human populations (S1 Table).

Banke District is a part of Lumbini Province located in mid-western Nepal with Nepalgunj as its District headquarter. It has an area of 2,337 sq. km elevated from 129 to 1,290 masl across a longitude of 81°29’ to 82°87’ East and latitude of 27°51’ to 28°20’ North. The temperature ranges from 4–46 °C in winter and summer, respectively [29]. The total number of households in this District are 129,307 representing 603,194 human populations (S1 Table).

#### Study populations

The study populations were sheep and goats reared in the aforementioned districts. The total sheep and goat populations in Jumla, Surkhet and Banke Districts were 73,241 and 44,677; 5,892 and 334,720; 9,700 and 322,897, respectively [30] (S1 Table). In Jumla, sheep and goats were mainly kept under transhumance production systems whereas in Surkhet and Banke Districts, extensive and semi-intensive farming systems were common (some farmers rear animals intensively, too).

### Study design and sample size

We carried out a cross-sectional study in the farms selected conveniently based on farmers’ willingness and accessibility from November 2023 to June 2024. We calculated the sample size by using Cochran’s formula based on the expected prevalence of 36.99% pooled from [25,31,32] with precision of 5% and 95 % of confidence level. The calculated required sample size (n) was 358. However, total number of sampled animals was increased to 368 for better inclusion and accuracy. Out of total samples, 107 from Jumla, 136 from Surkhet and 125 from Banke were sampled from live animals from farms.

### Sample collection and serological examination

We collected a total of 5 ml of blood from the jugular vein of the selected animals (S1 Figure) and transferred each of these blood samples into sterile plain tubes and stored in an icebox until transported them to the Veterinary Laboratory, Surkhet under the Ministry of Agriculture and Livestock Development. Then, we separated sera from the respective blood samples using a centrifuge (1,500 rpm for 15 min) and stored these sera at −20 °C until the indirect enzyme-linked immunosorbent assay (iELISA) test was performed to detect anti-*T. gondii* antibodies (IgG) (S2 Figure).

We performed the iELISA test with wells coated with P30 *T. gondii* antigen using ID Screen Toxoplasmosis indirect multi-species (TOXOS-MS-2P, ID Vet, France) to detect anti-Toxoplasma antibodies according to the manufacturer’s procedure (Appendix1) (S3 Figure).

### Questionnaire survey

We interviewed farm attendants from each farm included in this study using a structured questionnaire at respective farms to identify risk factors of *T. gondii* infection in sheep and goats. We assessed herd size, demographics (age, sex, location of farms), altitudinal gradient, management practices, cat presence/absence in and around farms and houses as well as their access to grazing lands, feed sources and water sources of sheep and goats and reproductive history (pregnancy and abortion) of the farmed sheep and goats (Appendix2). Further, we identified risk of *T. gondii* infection to the farmers and their family members by evaluating additional factors such as drinking water treatment and habit of eating raw or undercooked meat and drinking raw or unpasteurized milk of sheep and goats. Additionally, we asked farm attendants to assess their knowledge on zoonotic diseases including toxoplasmosis (S4 Figure).

### Data analysis

After data coding, we entered data and laboratory-based information in a Microsoft Excel spreadsheet and analyzed these data using IBM SPSS Statistics 26 (SPSS Inc., IBM Corporation, Version 26, Chicago, IL, USA). We estimated the seroprevalence by dividing the number of seropositive samples by the total samples. We considered sheep and goats from a single farm as a single herd also considered positive herd having at lease one animal seropositive. We determined herd and individual animal-wise seroprevalence. We categorized herd size as less than 30 and more than 30, respectively as small and large herds. We also categorized age according to those having less than 12 months, 12 to 36 months and more than 36 months.

We scored and ranked the behaviours and knowledge of farm attendants that would keep them at risk of *T. gondii* infection and measured the correlation (r) between seroprevalence in herds and the total risk index using Spearman’s rank correlation. We ranked farm attendants having 0% risk behaviours as ‘no risk at all’, 1–19% as ‘negligibly vulnerable’, 20–59% as ‘moderately vulnerable’ and 60–100% as ‘highly vulnerable’ to *T. gondii* infection. Similarly, we applied spearsman’s rank correlation to determine the relation between elevation and herd seroprevalence. All statistical tests were made at 5% significance level and p-values less than 0.05.

## Results

### Seroprevalence of *T. gondii* among herds

The blood samples were collected from 69 herds of sheep and goats reared in Jumla (17.4%), Surkhet (49.3%) and Banke (33.3%) Districts (Table 1). At least one sheep or goat was infected by *T. gondii* in 64 out of 69 herds [Median = 12, IQR (Q_3_-Q_1_) = 10, minimum = 2, maximum = 150] included in our study suggesting 92.8% overall seroprevalence. The median prevalence of *T. gondii* among 64 herds was 70% (IQR (Q_3_-Q_1_) = 73.22, minimum = 2, maximum = 150, S2 Table).

**Table 1.** Seroprevalence and risk analysis associated with farm characteristics of herds.

| Farm characteristics | | Total sample | Percentage | Number of positive herds | Seroprevalence % | $\chi^2$ value | df | p-value |
| --- | --- | --- | --- | --- | --- | --- | --- | --- |
| Location | Jumla (3522–3578 m asl) | 12 | 17.4 | 11 | 91.7 | 0.434 | 2 | 0.81 |
|  | Surkhet (481–675 m asl) | 34 | 49.3 | 31 | 91.2 |  |  |  |
|  | Banke (169–174 m asl) | 23 | 33.3 | 22 | 95.7 |  |  |  |
| Overall |  | 69 |  | 64 | 92.75 |  |  |  |
| Herd size | upto 30 | 56 | 81.2 | 51 | 91.10 | 0.020 | 1 | 0.68 |
|  | more than 30 | 13 | 18.8 | 12 | 92.31 |  |  |  |
| Rearing | Extensive | 12 | 17.4 | 11 | 91.67 | 2.131 | 2 | 0.35 |
| system | Semi-intensive | 45 | 65.2 | 43 | 95.56 |  |  |  |
|  | Intensive | 12 | 17.4 | 10 | 83.33 |  |  |  |
| Types of animals reared | Sheep | 27 | 39.1 | 25 | 92.59 | 1.795 | 3 | 0.41 |
|  | Goat | 31 | 44.9 | 27 | 87.10 |  |  |  |
|  | Mixed | 11 | 15.9 | 11 | 100.00 |  |  |  |
| Hygienic condition | Good | 14 | 20.3 | 12 | 85.71 | 1.435 | 2 | 0.49 |
|  | Fair | 52 | 75.4 | 49 | 94.23 |  |  |  |
|  | Poor | 3 | 4.3 | 3 | 100.00 |  |  |  |
| Presence of domestic cats in and around farms and houses | Yes | 8 | 11.6 | 8 | 100 | 0.707 | 1 | 0.40 |
|  | No | 61 | 88.4 | 56 | 91.80 |  |  |  |
| Presence of feral/wild cats | Yes | 60 | 87.0 | 55 | 91.67 | 0.809 | 1 | 0.37 |
|  | No | 9 | 13.0 | 9 | 100.00 |  |  |  |
| Access of cats to grazing land | Yes | 18 | 26.1 | 17 | 94.44 | 0.104 | 1 | 0.75 |
|  | No | 51 | 73.9 | 47 | 92.16 |  |  |  |
| Access of cats to animal feeds | Yes | 33 | 47.8 | 30 | 90.91 | 0.320 | 1 | 0.57 |
|  | No | 36 | 52.2 | 34 | 94.44 |  |  |  |
| Access of cats to water source | Yes | 19 | 27.5 | 16 | 84.21 | 2.847 | 1 | 0.09 |
|  | No | 50 | 72.5 | 48 | 96.00 |  |  |  |

The study farmers distributed across 577 m median elevation asl (IQR (Q_3_-Q_1_) = 500 m asl; minimum = 169, maximum = 3578 m asl). This elevation was positively and weakly correlated with herd-level seroprevalence (S3 Table).

### Seroprevalence of *T. gondii* among individual-level of sheep and goats

Among 368 serum samples examined, we detected *T. gondii* antibodies in 227 samples (61.7%); 81.4% of sheep (n = 136/167) and 45.3% of goats (n = 91/201) were found seropositive which was highly associated (p < 0.001) suggesting sheep being more vulnerable to *T gondii* infection (Table 2).

**Table 2.** Seroprevalence and risk analysis associated with locations and biological characteristics of sampled animals.

| Variables | Categories | Total sample | Percentage | No. of positive sample | Seroprevalence % | $\chi^2$ - value | df | p-value |
| --- | --- | --- | --- | --- | --- | --- | --- | --- |
| Location (Districts) | Jumla | 107 | 0.0 | 91 | 85.0 | 38.145 | 2 | 0.012 |
|  | Surkhet | 136 | 37.0 | 78 | 57.3 |  |  |  |
|  | Banke | 125 | 34.0 | 58 | 46.4 |  |  |  |
| Overall |  | 368 |  | 227 | 61.7 |  |  |  |
| Host animal | Sheep | 167 | 45.38 | 136 | 81.4 | 50.473 | 1 | <0.001 |
|  | Goat | 201 | 54.62 | 91 | 45.3 |  |  |  |
| Age (months) | <12 | 161 | 43.8 | 87 | 54.0 | 9.545 | 2 | <0.001 |
|  | 12–36 | 132 | 35.9 | 84 | 63.6 |  |  |  |
|  | >36 | 75 | 20.4 | 56 | 74.7 |  |  |  |
| Sex | Male | 131 | 35.6 | 79 | 60.3 | 0.164 | 1 | 0.082 |
|  | Female | 237 | 64.4 | 148 | 62.4 |  |  |  |
| Pregnancy | Yes | 26 | 7.1 | 15 | 57.7 | 0.282 | 1 | 0.370 |
|  | No | 211 | 57.3 | 133 | 63.0 |  |  |  |
| History of abortion | Yes | 25 | 6.8 | 19 | 76.0 | 2.189 | 1 | 0.102 |
|  | No | 212 | 57.6 | 129 | 60.9 |  |  |  |

### Risk factors

We found altitude, age of animal, type of host animal, herd size, rearing system, types of animals reared in a herd, presence of domestic cats and access of cats to water sources as risk factors. The highest prevalence was found in Jumla (higher land) followed by Surkhet (mid land) and Banke (low land) Districts (p = 0.012). Among the different age groups, the highest seroprevalence was in >36 months (74.7%) followed by 12–36 months (63.6%) and <12 months (54%) (p <0.001). Larger herds were mostly affected (83.9%) by *T. gondii* infection than small herds (52%) (p <0 .001). Extensive rearing systems had the highest impact (85%) while intensive (71.4%) and semi-intensive rearing system (50.4%) had a lesser impact on seroprevalence of *T. gondii* (p < 0.001). Rearing of only sheep in the farm had higher rate of *T. gondii* infection (82.4%) followed by the mixed herd type (77.8%) and the herds having only goats (39.5%) (p < 0.001). Next, 39.1% of sheep and goats reared along with domesticated cats showed seropositivity. Among animals where cats had access to the drinking water sources, 49% were found positive (Table 3).

**Table 3.**
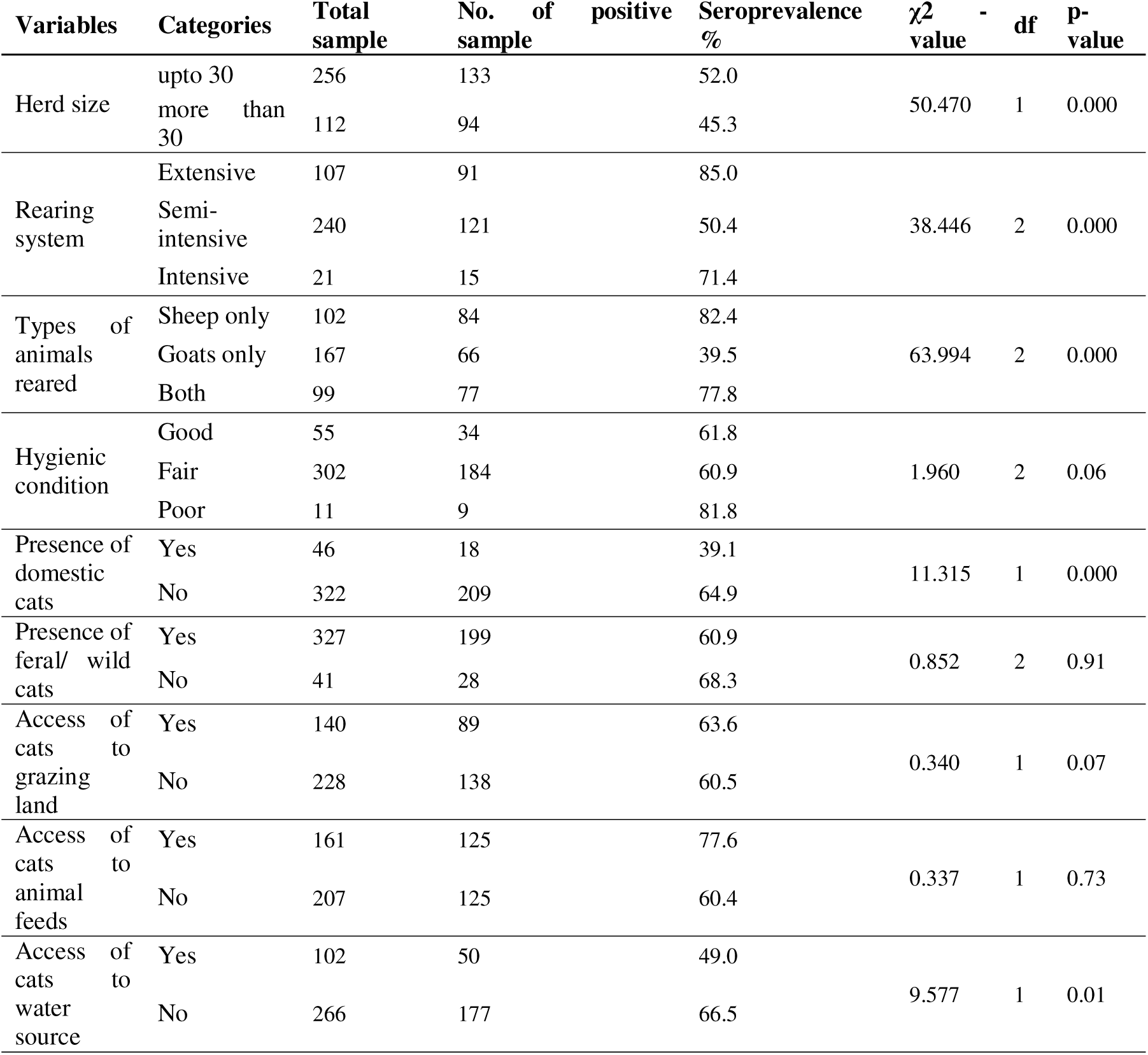
Seroprevalence and risk analysis associated with farm characteristics of sheep and goats.

### Risk of *T. gondii* infection to farm attendants and their knowledge of zoonotic diseases including toxoplasmosis

Among 69 farm attendants interviewed during the study, a total of 83% of farmers included in this study had moderate risk behaviors which kept them at risk of being infected by *T. gondii* (Table 4). A moderate correlation (r = 0.5) was found between these risky behaviours of farm attendants and seroprevalence of *T. gondii* in sheep and goat herds (S3 Table).

**Table 4.** Characteristics and knowledge of farm attendants on zoonotic disease and toxoplasmosis.

| Variables | Categories | Frequency | Percentage |
| --- | --- | --- | --- |
| Habit of eating raw/ undercooked meat | Yes | 56 | 81.2 |
|  | No | 13 | 18.8 |
| Habit of drinking raw/ unpasteurized milk | Yes | 0 | 0 |
|  | No | 69 | 100 |
| Treatment of water before drinking | Yes | 10 | 14.5 |
|  | No | 59 | 85.5 |
| Knowledge on zoonotic disease | Yes | 35 | 50.7 |
|  | Rabies | 23 | 65.71 |
|  | Parasite | 2 | 5.71 |
|  | Rabies, Parasite | 1 | 2.86 |
|  | Rabies, Birdflu | 8 | 22.86 |
|  | Rabies, Corona | 1 | 2.86 |
|  | Jumla | 5 | 14.29 |
|  | Surkhet | 17 | 48.57 |
|  | Banke | 13 | 37.14 |
|  | No | 34 | 49.3 |
| Knowledge on Toxoplasmosis | Yes | 0 | 0 |
|  | No | 69 | 100 |
| Risk ranking (Vulnerability) | Percent of risk index |  |  |
| No risk at all | 0% | 7 | 10.14 |
| Negligible | 1–19% | 0 | 0 |
| Moderate | 20–59% | 57 | 82.61 |
| High | 60–100% | 5 | 7.25 |

## Discussion

Our study had revealed the widespread of *T. gondii* in sheep and goats reared in mid-western part of Nepal.

### Seroprevalence among herds

We confirmed seropositivity among studied herds of sheep and goats reared across lowlands to highlands of mid-western Nepal. We found lower herd-level seroprevalence (92.8%) than the seropositivity recorded in herds of sheep and goats reared in Algeria (96.2%) [33] and Italy (97%) [34]. However, our results were slightly greater than those reported from sheep and goat herds reared in South Africa (88.3%) [35].This suggests widespread contamination of the *T. gondii* oocysts in animals’ farms and their grazing lands, feeds and water sources. Commonly distributed domestic and feral cats (12–87%) in and around residences and animal farms, sanitation, environmental conditions might be responsible for the dissemination of these oocysts in those environments which put both animals and farm attendants are at moderate risk for acquiring *T. gondii* infection (r = 0.5).

### Seroprevalence at the individual level and risky biological characteristics of animals

The overall seroprevalence of *T. gondii* infection in our study was higher (61.7%), than the finding reported from a similar study carried in 34 different districts of Nepal (14.6%) [22]. Other similar studies conducted elsewhere worldwide also had lower seroprevalence [13.9% in South Africa [36], 50.5% in Italy [37], and 32.3% in Pakistan [38]]. In the neighboring country India, prevalence in sheep and goats ranges from 1.4% to 42.5% [31,39]. The seropositivity of our study was almost similar to the seropositivity of *T. gondii* (62.8%) reported from sheep and goats farmed in Romania [40]. A higher rate of prevalence was observed in various studies either with multiple species or single species. In Sao Tome and Principe, 68.4% of sheep and 70.1% of goats were detected as seropositive to *T. gondii* [41]. The differences in the seroprevalence observed at individual levels between our study and aforementioned studies could be due to the access of sheep and goats to contaminated feeds and water, the agro-climatic variations, the age and sex of examined animals, the diagnostic and sampling techniques used, owner’s knowledge on zoonotic diseases and toxoplasmosis, rearing system and animal hygiene. The higher seroprevalence in present study could be ascribed to suitable climatic conditions and the widespread presence of cats.

#### Location wise seroprevalence

Our study determined altitude as a risk factor of *T. gondii* in sheep and goats of study area. The overall seroprevalence of *T. gondii* at the individual animal level was highest in Jumla District (85%) as compared to Surkhet (57.3%) and Banke (46.4%) Districts. Our finding is in consistent with that of China where high seroprevalence of *T. gondii* was detected in Tibetian sheep in the Gansu province of which was at more than 1000 meters above sea level [42]. Our study also agrees with the finding from Ethiopia where sheep and goats from higher altitude had highest seroprevalence followed by midland and low land [43]. The animal rearing system in Jumla was based on a transhumance migratory system (Extensive System) and the study animals were from rural area and and have longer grazing time in poorly managed pasture land. Even though the presence of domestic cats was less, contamination of pasture by wild cats was likely to happen as the grazing land was near to the forest. At least a dog was reared by the farmers in Jumla for security purposes which may act as vector for *T. gondii* transmission [44]. All of these factors may be the contributor for the highest seroprevalence of *T. gondii* in Jumla District. In Surkhet and Banke Districts, lower seroprevalence may be because of better veterinary services, rearing systems and awareness of people about zoonotic diseases.

#### Host animal wise seroprevalence

Sheep had significantly higher seroprevalence of *T. gondii* (81.4%) as compared to goats (45.3%) in our study. This agrees with findings of other studies that sheep are more prone to toxoplasmosis than goats as in Nepal [22], India [45], and Ethiopia [46] however other studies are in contrast to this findings and have a higher prevalence in goats than in sheep as in and Pakistan [38]. Rearing of sheep alone in the herd had higher seroprevalence than rearing goats alone and mixing of both sheep and goats in our study. The difference in seroprevalence between the sheep and goats could be due to eating habits. Goats are browsers that prefer leaves and twigs from taller bushes and shrubs while sheep are grazers who eat short grasses near the soil [47] making them more prone to get infected by *T. gondii*. In our study area, sheep were mostly reared in higher land where animals were kept under extensive system which may be contribute to the result.

Among sheep, the result of our study is greater than 36.2% in Nepal [25], and 50.6% in Romania [40] and 33.7% in Ethiopia [46]. Seroprevalence in sheep was higher in Serbia [48] (84.5%) in and almost equal to 80% in Brazil [49]. In case of goats, a higher prevalence was observed; 52.2% in Saudi Arabia [50] and 53.8% in Pakistan [38]. than in our findings. The result of our study is in contrast with findings of others as 96% in Nepal [23] and 13% in China [51]. Seroprevalence in goats of the present study is almost similar to that of India (42.4%) [31]. In a recent study conducted among the aborted ewes of Iran, 61.5% were serodiagnosed with toxoplasmosis [52]. Beside this, many sheep and goats slaughtered for human consumption were found infected by *T. gondii* [20]. The differences in the seroprevalence of *T. gondii* in sheep and goats in different parts of the world might be due to the difference in geo-climatic conditions, altitude, animal rearing system, and knowledge of farmers on zoonotic diseases including toxoplasmosis.

#### Seroprevalence and demographics of sheep and goat individuals

We detected higher seroprevalence in the older age group (i.e., >36 months) of sheep and goats than in adults and young ones. This result agrees with many other studies; in Nepal [23] and Romania [40]but in Libia, seroprevalence was higher in younger sheep and goats [53]. This difference in seropositivity among different age groups of animals may be due to higher chances of acquiring infection by older animal over a long period.

### Seroprevalence and risk analysis associated with farm characteristics of animals

We detected herd size, rearing system, types of animals reared, presence of domestic cats, access of cats to water sources as the significant risk factors associated to the infection of *T. gondii* in sheep and goats. Among the different herd size, larger herds had higher chances of getting an infection in our study. Findings fromPakistan [54] agrees with our findings. Eventhough the *T. gondii* infection do not transfer between animals but can be congenital [55]. The large herd contains offspring from the parent sheep and goats of the same herd. Higher seroprevalence in larger herds might be due to the fact that larger herds in the study area were reared under extensive system which increases the chances of acquiring toxoplasmosis as compared to animals reared under other systems.

An extensive system of rearing animals has the highest impact on the occurrence of toxoplasmosis in sheep and goats in our study. This agrees with the previous study conducted in China [56] but in contrast to the result of Brazil showing that semi-intensive system of rearing had the highest seroprevalence rate than in extensive and intensive system [57]. Sheep and goats under the extensive system were grazed on pasture which possess a risk of getting oocyst shed by wild, stray or domestic cats while in the intensive management system, animals were offered feed and water within the shed and get less chances to ingest oocyst.

Cats are the definitive host of *T. gondii*. Other animals acquire by ingesting oocyst from contaminated feed, water and soil. Oocyst present in the feces of cats was found in different parts of the world [58] and also from the serum samples [59]. So, the presence of cats either domestic, feral or wild roaming in and around house and farm and also their access to water, feed, pasture, vegetable garden can transmit oocyst to animals and humans. Other studies have also associated the presence of domestic cats as a risk factors for *T. gondii* [60]. But in our study, presence of cats (n=8) had less influence in the seroprevalence of *T. gondii.* In the absence of domestic cats, stray or feral cats may be less monitored or controlled and may serve as alternative sources of oocyst contamination [61]. This environmental persistence of *T. gondii* oocysts means that herds can still become infected through indirect transmission routes even without direct contact with infected cat feces.

Water contamination by the oocyst of *T. gondii* shed by the cats in the feces is one of the routes of transmission to humans and animals [62]. In this study, sheep and goats drinking water from sources that was not accessed by cats were found high seropositive as compared to sheep and goats that drank water from sources that were accessed by cats. This suggests that the water is less contaminated by the oocyst of *T. gondii*.

### Human risk factors

There were some study carried in human toxoplasmosis in Nepal [63–67]. First case of congenital toxoplasmosis in 53 days old human baby from Nepal was reported for the first time in 2011 [68]. There are reports of seroprevalence of *T. gondii* detected among HIV/AIDS patients in Kathmandu [64] and among pneumonic cases known in Kathmandu, Nepal [69].

Our survey indicated that half of the respondents had no knowledge and none of them were aware of toxoplasmosis. Those who were aware of zoonoses had only heard about rabies, bird flu, coronavirus and parasites but not about the specific parasite. A study conducted on the knowledge and practices of livestock farmers in three different Districts of Nepal (Tanahu, Manang and Nawalpur), revealed that about 95% knew about birdflu while 90.7% knew about rabies [70]. A survey in Ethiopia revealed that 52.5% of respondents knew about zoonotic disease. Among them, about 90% were familiar with rabies. More than half of the respondents had a habit of eating raw meat and drinking raw milk [71]. These factors contribute to the transmission of different zoonotic diseases including toxoplasmosis. Also, most of the respondents had a habit of eating undercooked or raw chevon and mutton in our study. A study conducted in 1998 A.D. in humans of Nepal detected high seroprevalence of toxoplasmosis associated with meat-eating habits [66]. Similarly, 33.3% of sheep and 32.5% of goats slaughtered for human consumption in Northwest Tunisia were infected by *T gondii* [72]. Similarly, 16% of sheep and goats in rural market in China [73] were infected. Cysts containing bradyzoites are present in the muscles, nerves and internal organs of sheep and goats. Eating raw or undercooked meat causes infection. More than one-third of respondents did not treat the water before drinking. Untreated or unfiltered drinking water can transmit the parasite if contamination has occurred [74]. Farm attandants and their family members did not practice drinking milk of sheep and goats in our study. In Nepal, goats are more important source of meat and easily digestible milk than sheep. Hence, goats are sometimes called as poor man’s cow [75]. This cultural practice may contribute to transmission of *T. gondii* among milk-feeding kids as well as consumers of raw goat milk in other parts of the country because Tavassoli [76] provided evidence of *T. gondii* presence in milk of sheep and goats of Iran and Sacks [77] mentioned similar infection patterns from these ruminants to the raw milk feeding humans in California.

Most of the farm attendants were at moderate risk for contracting toxoplasmosis based on their knowledge of zoonoses, meat-eating habits, drinking milk and water treatment practices (r = 0.5). Because of placental and breast-feeding transmission of toxoplasmosis in humans [78,79] and our findings of risky behaviour of farm attendats (Table 4), we suggest to conduct the sophisticated study in vulnerable human populations nationwide.

## Conclusion

The study has demonstrated that *T. gondii* infections are widely prevalent among sheep and goats in the mid-western region of Nepal. Identified risk factors for *T. gondii* include altitude, age of animal, type of host animal, herd size, rearing system, types of animals reared in a herd, presence of domestic cats and access of cats to water sources. The high seroprevalence of toxoplasmosis in sheep and goats indicates significant environmental contamination with *T. gondii* oocysts, largely due to the unrestricted access of cats to animal premises which has important public health implications. Therefore, it is essential to develop and implement preventive measures to address the risk factors and reduce both animal losses and zoonotic transmission to humans. Furthermore, future research should consider other common animals in the country and explore genetic-level study.

## Data Availability Statement

The data that support the findings of this study are available as supplementary files (S1 Data, S1–S4 Figures through S1–S3 Tables).

## S1 Data

Dataset used for quantitative analysis in this study.

## S1 Figure

Showing blood collection from sheep in Banke.

## S2 Figure

Showing the performance for ELISA test.

## S3 Figure

Showing ELISA test result.

## S4 Figure

Showing PI conducting a questionnaire survey.

*As the S1 Data and S1 to S4 figures (photographs of people including authors, research participants or volunteers, identifiable characteristics or settings (e.g., home environment) would allow the patient/study participants or their family, friends or neighbors to identify them (e.g., age, precise location, farm owner’s name, etc.), readers are humbly requested to contact with the corresponding author to access to these supplementary files*.

## Acknowledgments

We are greatful to Ministry of Land Management, Agriculture and Cooperatives, Karnali Province for research grant (program number: 31200909; program indication number: 2.7.15.933), Veterinary Laboratory, Surkhet and for laboratory facilities as well as all the lab attandants for technical help.

## Author Contributions

**Conceptualization:** Bhrikuti Bhattarai Sharma, Ananta Dahal, Deb Prasad Pandey.

**Data curation:** Bhrikuti Bhattarai Sharma, Deb Prasad Pandey.

**Formal analysis:** Bhrikuti Bhattarai Sharma, Jaya Prasad Singh.

**Funding acquisition:** Bhrikuti Bhattarai Sharma.

**Investigation:** Bhrikuti Bhattarai Sharma, Anup Adhikari, Prativa Shrestha.

**Methodology:** Bhrikuti Bhattarai Sharma, Deb Prasad Pandey, Jaya Prasad Singh.

**Project administration:** Bhrikuti Bhattarai Sharma, Ananta Dahal, Rebanta Kumar Bhattarai (facilitation for ethical approval).

**Resources:** Bhrikuti Bhattarai Sharma, Ananta Dahal, Anup Adhikari.

**Supervision:** Ananta Dahal, Deb Prasad Pandey.

**Validation:** Ananta Dahal, Deb Prasad Pandey.

**Visualization:** Bhrikuti Bhattarai Sharma, Deb Prasad Pandey.

**Writing – original draft:** Bhrikuti Bhattarai Sharma, Deb Prasad Pandey.

**Writing – review & editing:** Bhrikuti Bhattarai Sharma, Ananta Dahal, Deb Prasad Pandey, Rebanta Kumar Bhattarai, Prativa Shrestha, Anup Adhikari, Jaya Prasad Singh.

**Competing interests:** The authors have declared that no competing interests exist.

## Appendices

## Appendix 1. iELISA test protocol

1. Before the test begins, we homogenized all the reagents and samples by vertex and allowed to come to room temperature (21 °C).
2. Initially, we added 90 microliters of dilution buffer to each microwell. Then we added 10 microliters of negative control to A1 and B1, 10 microliters of positive control to C1 and D1 and 10 microliters of each serum sample to the remaining wells.
3. After that we covered the plate and incubated for 45 minutes at 21 °C. After emptying the wells, we washed each of wells 3 times with 300 microliters of wash solution (1X).
4. Then, we added 100 microliters of conjugate (1X) to each well and incubated for 30 minutes.
5. Again, after emptying the wells, we again washed each well three times by wash solution.
6. After that, we added 100 microliters of substrate solution to each well and incubated for 15 minutes at 21 °C in the dark.
7. At last, we added 100 microliters of stop solution to each well to stop the reaction. In the presence of antibodies, blue color changed to yellow.
8. Then we placed the plate in the ELISA microplate reader and recorded Optical Density (O.D.) at 450 nanometres (nm).

## Appendix 2. Questionnaire for assessing risk factors

### Farm Characteristics

1. Date of interview…………………………………………………………………
2. Name of interviewee ………………………………………….…………………
3. Address ………………………………………………………………………
4. Species reared: (i) Sheep (ii) Goats (iii) Both
5. Herd size…………………………………………………. ……
6. Farming activities: (i) Livestock (ii) Agriculture (iii) Mixed farming
7. Type of management system: (i) Semi-intensive (ii) Intensive (iii) Extensive
8. Do you have cats? ……………………………………………..
9. Do the cats have their own house? …………………………………………………………………………….
10. Is there any stray/wild cats visiting your site?…………………………………………………
11. Do these cats access the water sources where sheep/goats drink water? Yes/No…………………
12. Do these cats access the feeds stored for sheep/goats? Yes/No……………………
13. Do these cats access the pasture land where sheep/goats graze? Yes/No…………….
14. Do the animals (sheep/goat/cat) have access to the vegetable garden? Yes/No…………..
15. Hygienic condition of farm: (i) Poor (ii) Fair (iii) Good
16. Do you also see rats (and other Rodents) roaming around your house? …………………………..

### Characteristics and knowledge of farm attendants on zoonoses and toxoplasmosis

17 Do you drink milk of sheep and goat? Yes/No……………………………………….
18 Do you boil the milk before drinking? Yes/No…………………………………………….
19 Do you eat meat? Yes/No…………………………………………………………….
20 Does the meat you consume cook thoroughly? Yes/No…………………………………….
21 Have you ever eaten undercooked/raw meat of sheep and goat? Yes/No……………
22 How do you treat water before consumption? (i)Boiling (ii) Apply disinfectant (…), (iii) Filtering (iv) Not boiling/not treating
23 Have you ever consumed unwashed fruits and raw vegetables? Yes/No………………….
24 Are you aware of any disease, which can be transmitted from animals to human?……..
25 Mention the diseases……………………………………………………
26 Regarding toxoplasmosis, do you know this disease? Yes/No……………………..
27 Are you aware that toxoplasmosis can affect human? Yes /No……………………………..
28 Information on animals

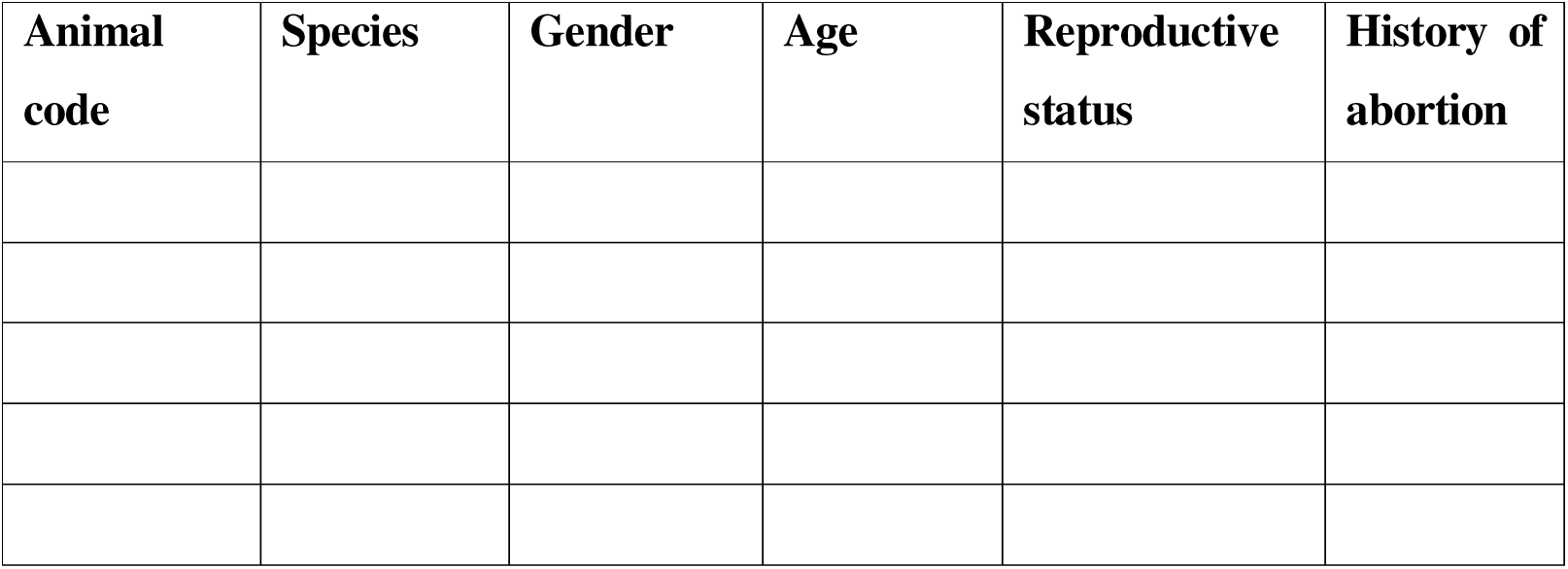

## Supplementary files

The supplementary files include S1 Data, and S1–S3 Tables, respectively. These are listed below.

**S1 Data: Dataset used for quantitative analysis in this study.**

Please, find the separately upoaded supplementary file named as “S1 Data_Dataset used for quantitative analysis in this study.xlsx”.

**S1 Table:** Human populations, sub-districts, wards and total populations of goats and sheep in districts selected for this study.

| Province | Karnali |  | Lumbini | Total |
| --- | --- | --- | --- | --- |
| Districts | Jumla | Surkhet | Banke |  |
| Human populations' | 118349 | 415126 | 60319 | 593794 |
| Households' | 24438 | 97893 | 129307 | 251638 |
| Total populations of sheep'' | 73,241 | 5,892 | 9,700 | 88,833 |
| Total populations of goat'' | 44,677 | 334,720 | 322,897 | 702,294 |
| Total sheep and goat populations | 117,918 | 340,612 | 332,597 | 791,127 |
| Total subdistricts | 8± | 9±± | 8±±± | 25 |
| Total selected subdistricts | 1 | 2 | 1 | 4 |
| *Represetation (in %) of subdistricts by this study* | 12.50 | 22.22 | 12.50 | 16 |
| Total wards in all subdistricts | 60± | 99±± | 81±±± | 240 |
| Total wards in selected subdistrict | 7 | 29 | 6 | 42 |
| Total selected wards | 1 | 3 | 1 | 5 |
| *Represetation (in %) of subdistricts by the wards* | 14.29 | 10.34 | 16.67 | 11.9 |
| Total subdivisions (i.e., toles/villages) of selected wards | 23^ | 56^^ | 8^^^ | 87 |
| Selected subdivisions in the wards | 2 | 4 | 1 | 7 |
| *Represetation (in %) of wards by toles /villages* | 8.70 | 7.14 | 12.50 | 8.04 |
| Total farms in the selected village | 250^ | 1645^^ | 400^^^ | 2295 |
| Total farms in the selected subdimision of the wards | 12 | 34 | 23 | 69 |
| **Total percent (%) of studied farms (i.e., ultimate study units)** | 4.8 | 2.06 | 5.75 | 3 |

**Symbols:** ±: https://daojumla.moha.gov.np/en/offices; ±±: https://daosurkhet.moha.gov.np/; ±±±: https://daobanke.moha.gov.np/; ' : National Statistical Office, 2021; " : https://www.dls.gov.np/downloadfiles/Livestock_Statistics_of_Nepal_2077_78_1659524236-1669717523.pdf; ^: Patarasi rural municipality; ^^: Birendranagar municipality ward number 10 office and Bheriganga municipality; ^^: Janaki rural municipality

**S2 Table:**
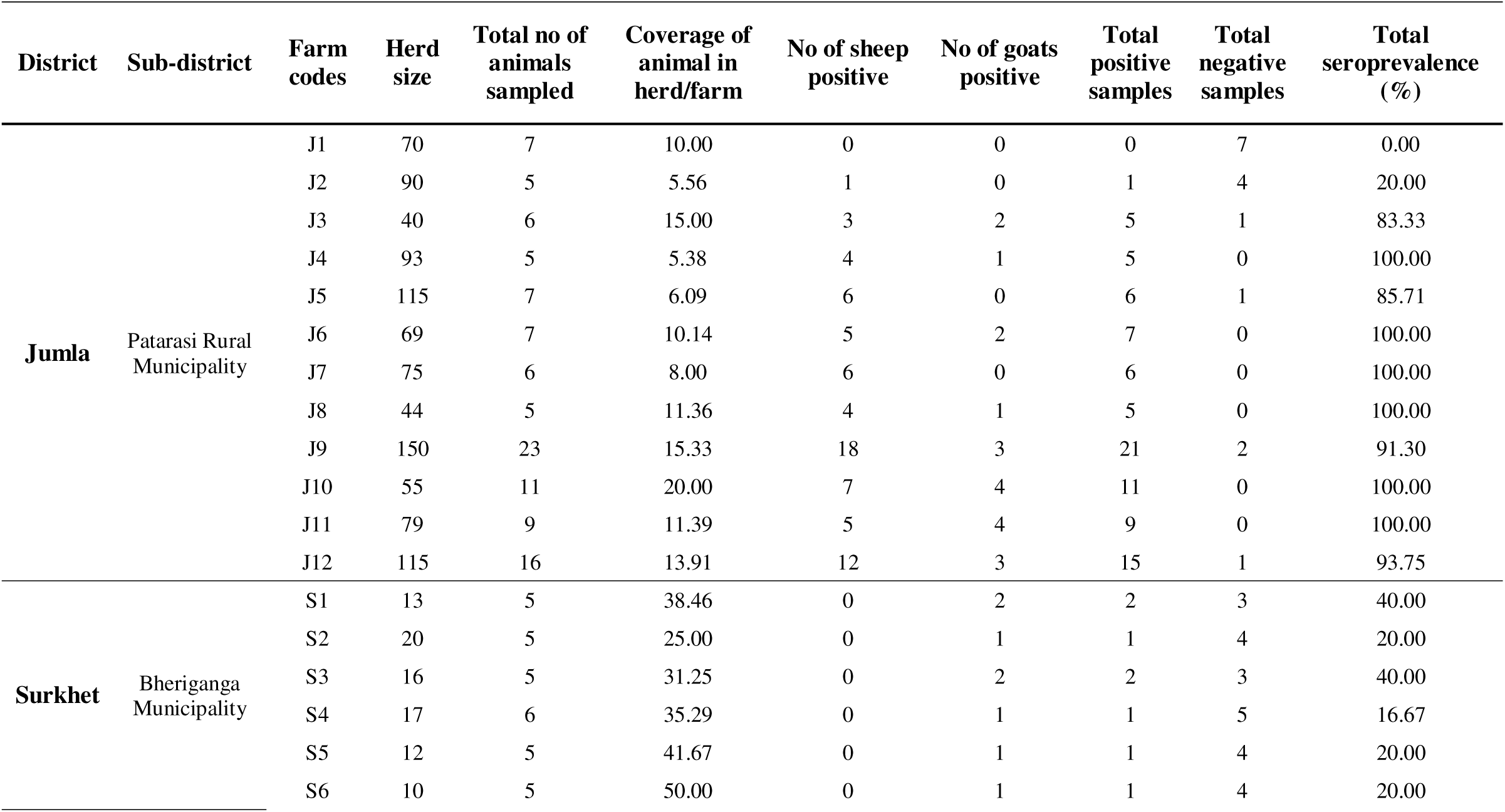
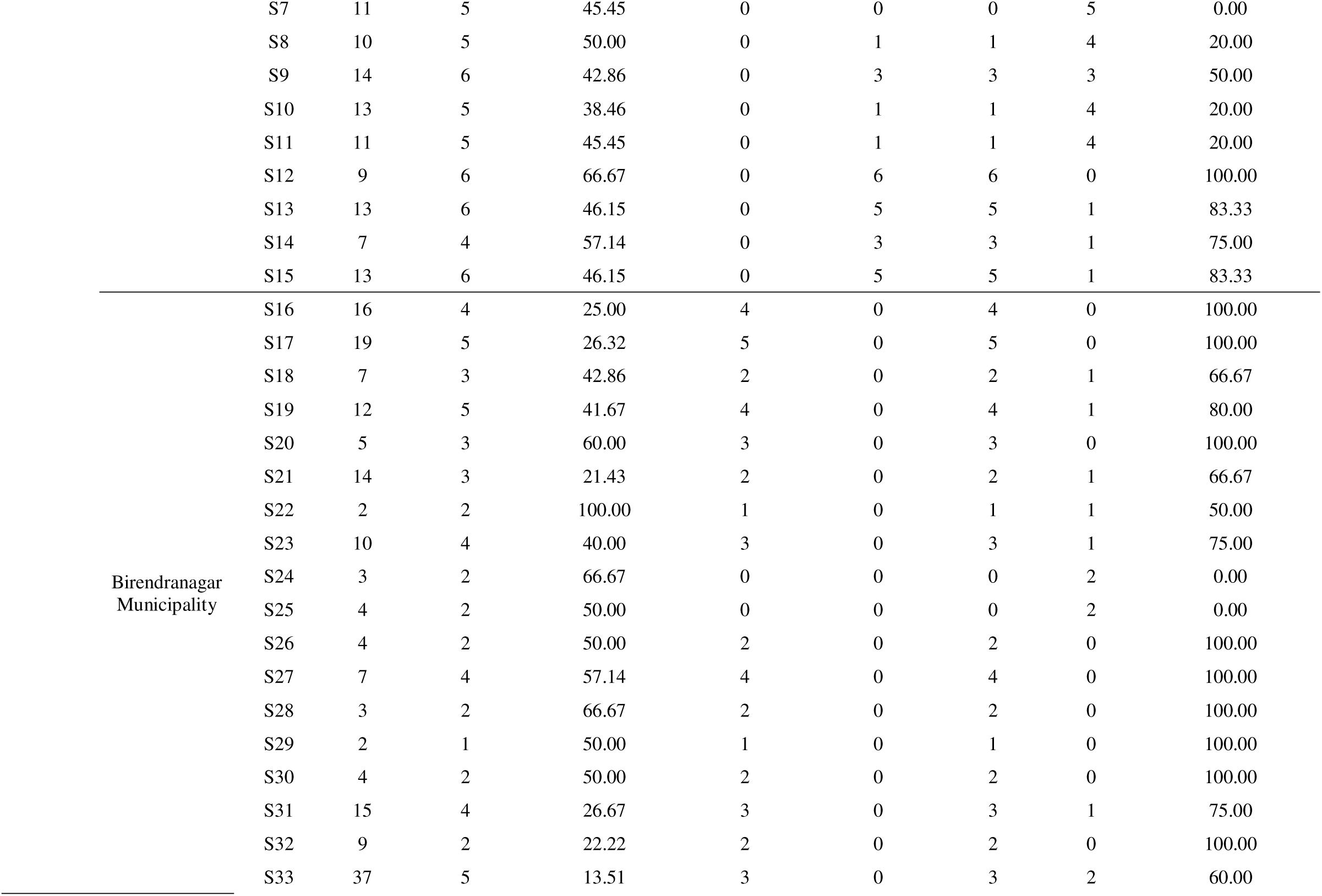
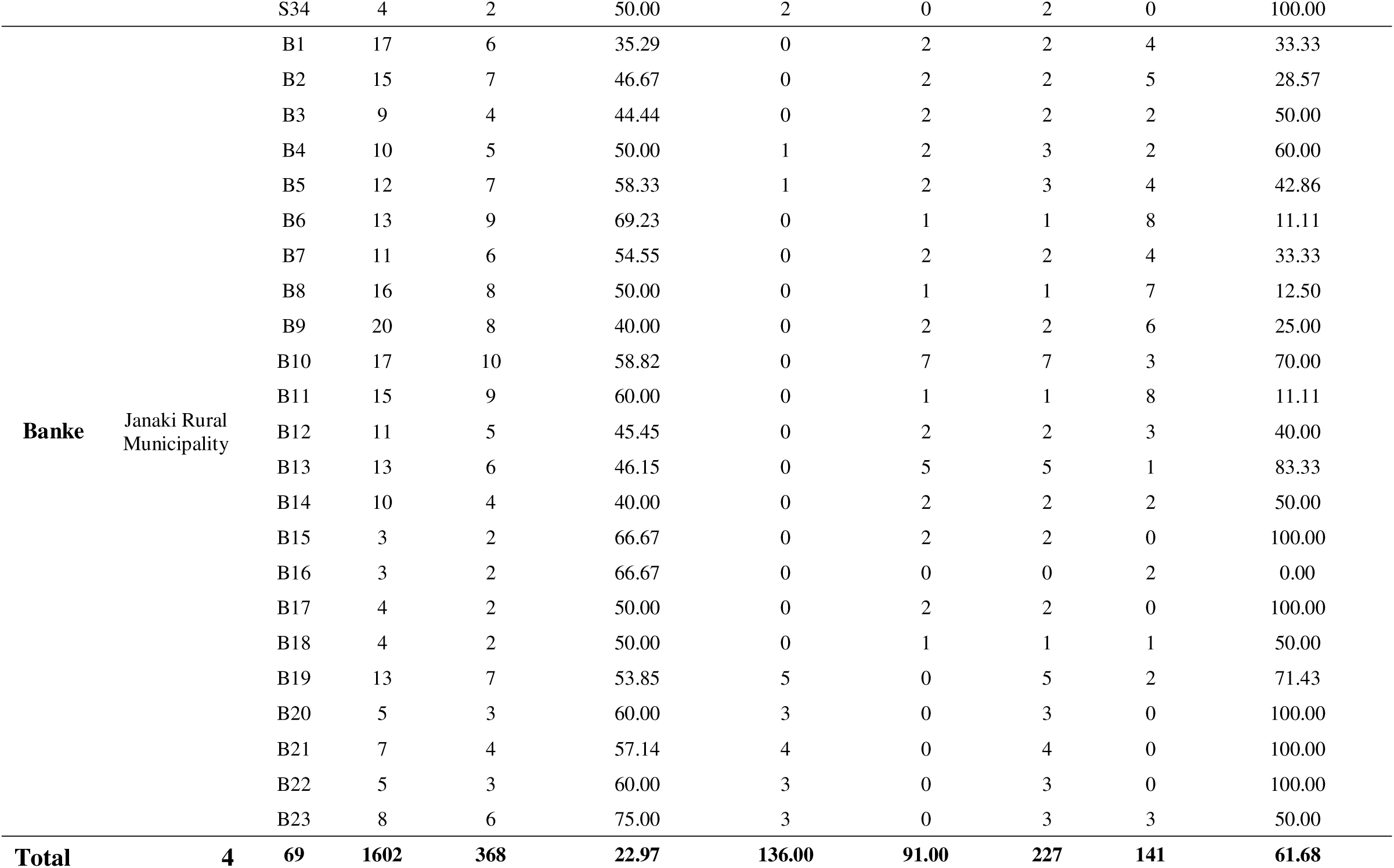
Locations of farms included in this study, size of herd and samples (sheep and goats) and seroprevalence of *T. gondii*.

**S3 Table.**
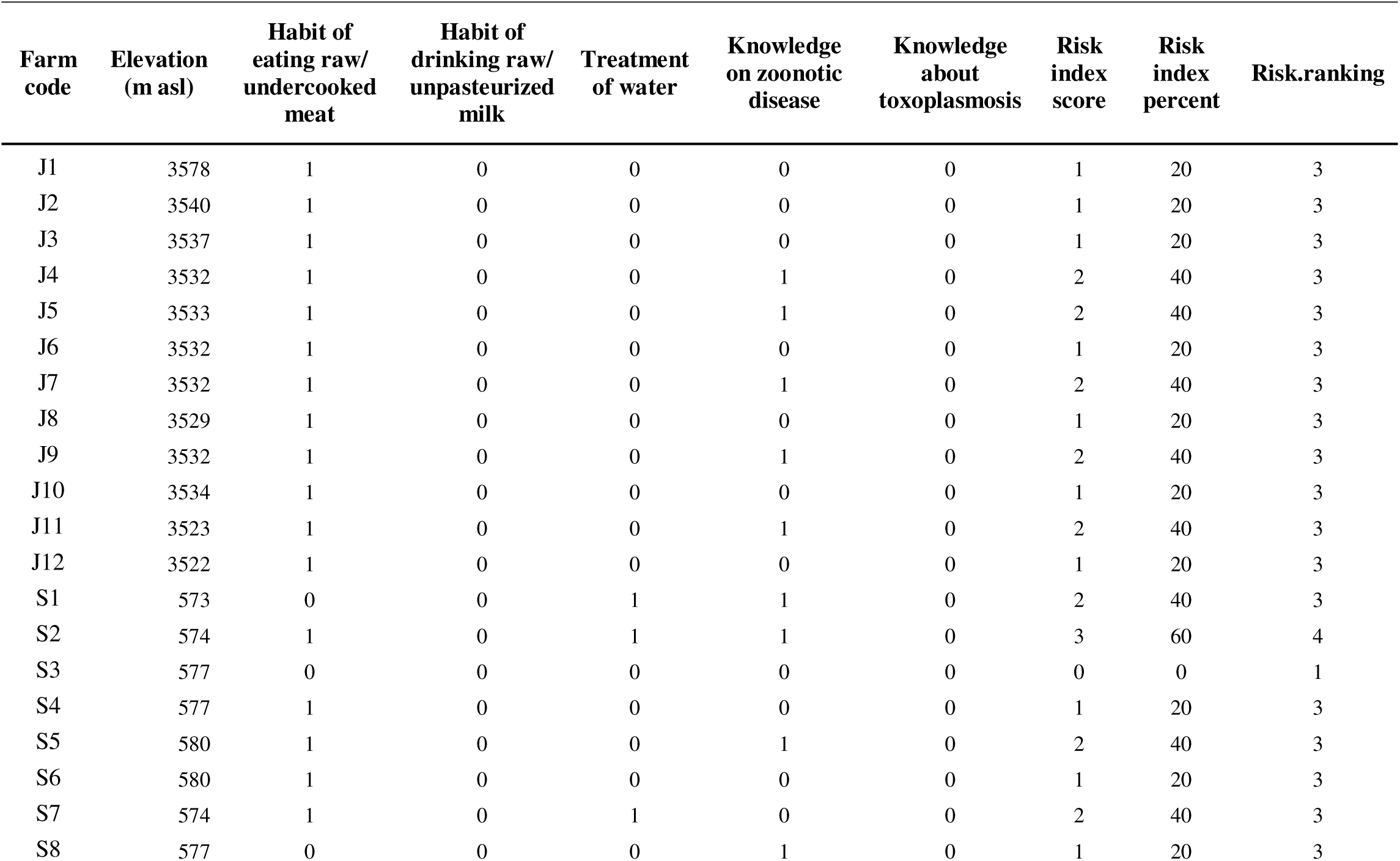
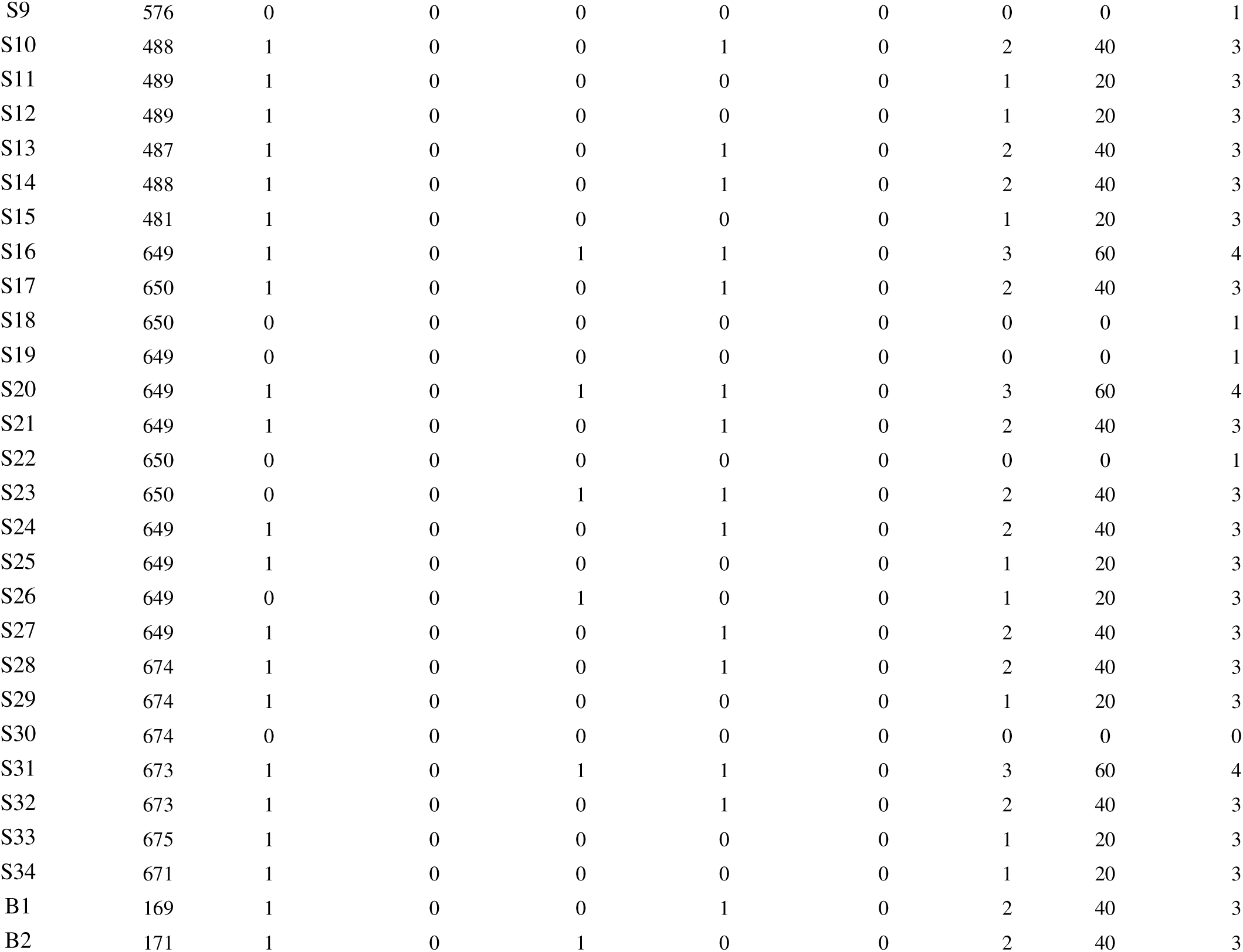
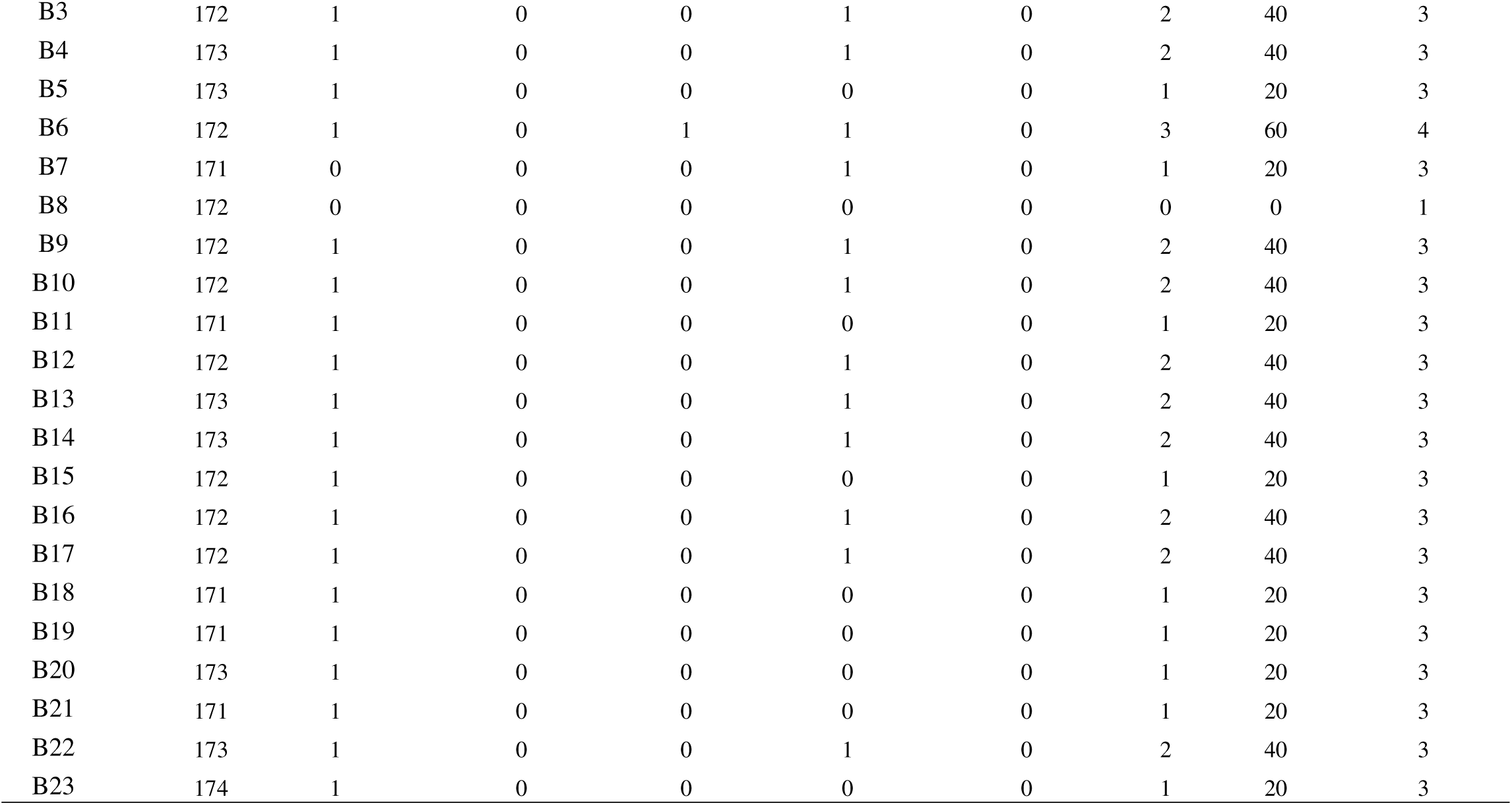
Human risk index.

| Farm code | Elevation (m asl) | Habit of eating raw/ undercooked meat | Habit of drinking raw/ unpasteurized milk | Treatment of water | Knowledge on zoonotic disease | Knowledge about toxoplasmosis | Risk index score | Risk index percent | Risk.ranking |
| --- | --- | --- | --- | --- | --- | --- | --- | --- | --- |
| J1 | 3578 | 1 | 0 | 0 | 0 | 0 | 1 | 20 | 3 |
| J2 | 3540 | 1 | 0 | 0 | 0 | 0 | 1 | 20 | 3 |
| J3 | 3537 | 1 | 0 | 0 | 0 | 0 | 1 | 20 | 3 |
| J4 | 3532 | 1 | 0 | 0 | 1 | 0 | 2 | 40 | 3 |
| J5 | 3533 | 1 | 0 | 0 | 1 | 0 | 2 | 40 | 3 |
| J6 | 3532 | 1 | 0 | 0 | 0 | 0 | 1 | 20 | 3 |
| J7 | 3532 | 1 | 0 | 0 | 1 | 0 | 2 | 40 | 3 |
| J8 | 3529 | 1 | 0 | 0 | 0 | 0 | 1 | 20 | 3 |
| J9 | 3532 | 1 | 0 | 0 | 1 | 0 | 2 | 40 | 3 |
| J10 | 3534 | 1 | 0 | 0 | 0 | 0 | 1 | 20 | 3 |
| J11 | 3523 | 1 | 0 | 0 | 1 | 0 | 2 | 40 | 3 |
| J12 | 3522 | 1 | 0 | 0 | 0 | 0 | 1 | 20 | 3 |
| S1 | 573 | 0 | 0 | 1 | 1 | 0 | 2 | 40 | 3 |
| S2 | 574 | 1 | 0 | 1 | 1 | 0 | 3 | 60 | 4 |
| S3 | 577 | 0 | 0 | 0 | 0 | 0 | 0 | 0 | 1 |
| S4 | 577 | 1 | 0 | 0 | 0 | 0 | 1 | 20 | 3 |
| S5 | 580 | 1 | 0 | 0 | 1 | 0 | 2 | 40 | 3 |
| S6 | 580 | 1 | 0 | 0 | 0 | 0 | 1 | 20 | 3 |
| S7 | 574 | 1 | 0 | 1 | 0 | 0 | 2 | 40 | 3 |
| S8 | 577 | 0 | 0 | 0 | 1 | 0 | 1 | 20 | 3 |
| S9 | 576 | 0 | 0 | 0 | 0 | 0 | 0 | 0 | 1 |
| S10 | 488 | 1 | 0 | 0 | 1 | 0 | 2 | 40 | 3 |
| S11 | 489 | 1 | 0 | 0 | 0 | 0 | 1 | 20 | 3 |
| S12 | 489 | 1 | 0 | 0 | 0 | 0 | 1 | 20 | 3 |
| S13 | 487 | 1 | 0 | 0 | 1 | 0 | 2 | 40 | 3 |
| S14 | 488 | 1 | 0 | 0 | 1 | 0 | 2 | 40 | 3 |
| S15 | 481 | 1 | 0 | 0 | 0 | 0 | 1 | 20 | 3 |
| S16 | 649 | 1 | 0 | 1 | 1 | 0 | 3 | 60 | 4 |
| S17 | 650 | 1 | 0 | 0 | 1 | 0 | 2 | 40 | 3 |
| S18 | 650 | 0 | 0 | 0 | 0 | 0 | 0 | 0 | 1 |
| S19 | 649 | 0 | 0 | 0 | 0 | 0 | 0 | 0 | 1 |
| S20 | 649 | 1 | 0 | 1 | 1 | 0 | 3 | 60 | 4 |
| S21 | 649 | 1 | 0 | 0 | 1 | 0 | 2 | 40 | 3 |
| S22 | 650 | 0 | 0 | 0 | 0 | 0 | 0 | 0 | 1 |
| S23 | 650 | 0 | 0 | 1 | 1 | 0 | 2 | 40 | 3 |
| S24 | 649 | 1 | 0 | 0 | 1 | 0 | 2 | 40 | 3 |
| S25 | 649 | 1 | 0 | 0 | 0 | 0 | 1 | 20 | 3 |
| S26 | 649 | 0 | 0 | 1 | 0 | 0 | 1 | 20 | 3 |
| S27 | 649 | 1 | 0 | 0 | 1 | 0 | 2 | 40 | 3 |
| S28 | 674 | 1 | 0 | 0 | 1 | 0 | 2 | 40 | 3 |
| S29 | 674 | 1 | 0 | 0 | 0 | 0 | 1 | 20 | 3 |
| S30 | 674 | 0 | 0 | 0 | 0 | 0 | 0 | 0 | 0 |
| S31 | 673 | 1 | 0 | 1 | 1 | 0 | 3 | 60 | 4 |
| S32 | 673 | 1 | 0 | 0 | 1 | 0 | 2 | 40 | 3 |
| S33 | 675 | 1 | 0 | 0 | 0 | 0 | 1 | 20 | 3 |
| S34 | 671 | 1 | 0 | 0 | 0 | 0 | 1 | 20 | 3 |
| B1 | 169 | 1 | 0 | 0 | 1 | 0 | 2 | 40 | 3 |
| B2 | 171 | 1 | 0 | 1 | 0 | 0 | 2 | 40 | 3 |
| B3 | 172 | 1 | 0 | 0 | 1 | 0 | 2 | 40 | 3 |
| B4 | 173 | 1 | 0 | 0 | 1 | 0 | 2 | 40 | 3 |
| B5 | 173 | 1 | 0 | 0 | 0 | 0 | 1 | 20 | 3 |
| B6 | 172 | 1 | 0 | 1 | 1 | 0 | 3 | 60 | 4 |
| B7 | 171 | 0 | 0 | 0 | 1 | 0 | 1 | 20 | 3 |
| B8 | 172 | 0 | 0 | 0 | 0 | 0 | 0 | 0 | 1 |
| B9 | 172 | 1 | 0 | 0 | 1 | 0 | 2 | 40 | 3 |
| B10 | 172 | 1 | 0 | 0 | 1 | 0 | 2 | 40 | 3 |
| B11 | 171 | 1 | 0 | 0 | 0 | 0 | 1 | 20 | 3 |
| B12 | 172 | 1 | 0 | 0 | 1 | 0 | 2 | 40 | 3 |
| B13 | 173 | 1 | 0 | 0 | 1 | 0 | 2 | 40 | 3 |
| B14 | 173 | 1 | 0 | 0 | 1 | 0 | 2 | 40 | 3 |
| B15 | 172 | 1 | 0 | 0 | 0 | 0 | 1 | 20 | 3 |
| B16 | 172 | 1 | 0 | 0 | 1 | 0 | 2 | 40 | 3 |
| B17 | 172 | 1 | 0 | 0 | 1 | 0 | 2 | 40 | 3 |
| B18 | 171 | 1 | 0 | 0 | 0 | 0 | 1 | 20 | 3 |
| B19 | 171 | 1 | 0 | 0 | 0 | 0 | 1 | 20 | 3 |
| B20 | 173 | 1 | 0 | 0 | 0 | 0 | 1 | 20 | 3 |
| B21 | 171 | 1 | 0 | 0 | 0 | 0 | 1 | 20 | 3 |
| B22 | 173 | 1 | 0 | 0 | 1 | 0 | 2 | 40 | 3 |
| B23 | 174 | 1 | 0 | 0 | 0 | 0 | 1 | 20 | 3 |

## Notes

### Competing Interest Statement

The authors have declared no competing interest.

### Author Declarations

This study was approved by the Institutional Review Board of AFU, Chitwan, Nepal (Ref. no. AFU-FW A00031653).

